# Altered glutamate signaling in Parkinson’s disease patients with REM sleep behavior disorder

**DOI:** 10.1101/2023.02.14.23285815

**Authors:** Christopher E.J. Doppler, Aline Seger, Ezequiel Farrher, Cláudia Régio Brambilla, Lukas Hensel, Christian P. Filss, Ana Gogishvili, N. Jon Shah, Christoph W. Lerche, Bernd Neumaier, Karl-Josef Langen, Gereon R. Fink, Michael Sommerauer

**Affiliations:** Cognitive Neuroscience, Institute of Neuroscience and Medicine (INM-3), Forschungszentrum Jülich, 52425 Jülich, Germany; University of Cologne, Faculty of Medicine and University Hospital Cologne, Department of Neurology, 50937 Köln, Germany; Institute of Neuroscience and Medicine (INM-4), Forschungszentrum Jülich, 52425 Jülich, Germany; Department of Nuclear Medicine, RWTH University Hospital, 52074 Aachen, Germany; Faculty of Medicine, RWTH Aachen University, 52056 Aachen, Germany; Engineering Physics Department, Georgian Technical University, Tbilisi, Georgia; Institute of Neuroscience and Medicine (INM-11), Molecular Neuroscience and Neuroimaging, JARA, Forschungszentrum Jülich, 52425 Jülich, Germany; JARA – BRAIN - Translational Medicine, 52056 Aachen, Germany; Department of Neurology, RWTH Aachen University, 52056 Aachen, Germany; Institute of Neuroscience and Medicine (INM-5), Forschungszentrum Jülich, 52425 Jülich, Germany

## Abstract

**Background and Objectives:** Clinical heterogeneity of patients with Parkinson’s disease is well recognized. Parkinson’s disease with rapid eye movement (REM) sleep behavior disorder (RBD) is a more malignant phenotype with faster motor progression and higher non-motor symptom burden. However, the neural mechanisms underlying this clinical divergence concerning disbalances in neurotransmitter systems remain elusive.

**Methods:** Combining magnetic resonance (MR) spectroscopy and ^11^C-ABP688 positron emission tomography (PET) on PET/MR hybrid system, we simultaneously investigated two different mechanisms of glutamate signaling in patients with Parkinson’s disease. Thirty-three patients were grouped according to their RBD status in overnight video-polysomnography and compared to 15 age- and sex-matched healthy control (HC) subjects. Total volumes of distribution (V_T_) of ^11^C-ABP688 were estimated with metabolite-corrected plasma concentrations during steady-state conditions between minutes 45 to 60 of the scan following a bolus-infusion protocol. Glutamate, glutamine, and glutathione levels were investigated with single voxel STEAM MR spectroscopy of the left putamen.

**Results:** We measured globally elevated V_T_ of ^11^C-ABP688 in patients with Parkinson’s disease and RBD compared to patients without RBD and HC subjects (*F*(2,45) = 5.579, *p* = 0.007). Conversely, glutamatergic metabolites did not differ between groups and did not correlate with the regional V_T_ of ^11^C-ABP688. V_T_ of ^11^C-ABP688 correlated with the amount of REM sleep without atonia (*F*(1,42) = 5.600, *p* = 0.023), and with dopaminergic treatment response in Parkinson’s disease patients (*F*(1,30) = 5.823, *p* = 0.022).

**Conclusion:** Our results suggest that patients with Parkinson’s disease and RBD exhibit altered glutamatergic signaling indicated by higher V_T_ of ^11^C-ABP688 despite unaffected glutamate metabolism. The disbalance of glutamate receptors and neurotransmitter might indicate a novel mechanism contributing to the heterogeneity of Parkinson’s disease and warrants further investigation of drugs targeting mGluR5.

## Introduction

Even though clinical diagnostic criteria of Parkinson’s disease are mainly focused on extrapyramidal motor impairment, considered to be primarily driven by dopaminergic depletion of the basal ganglia,^1^ clinical heterogeneity of the disease is well recognized.^2,3^ This resonates well with multiple neurotransmitters beyond the dopaminergic system being affected by misfolded α-synuclein, potentially implying differential neurotransmitter pathology in PD subtypes.^4,5^

Parkinson’s disease with rapid eye movement (REM) sleep behavior disorder (RBD) – a parasomnia characterized by insufficient muscle atonia and dream enacting behavior – is reported to exhibit a more malignant phenotype with faster motor progression and a higher burden of non-motor symptoms.^6,7^ Using positron emission tomography (PET), profuse demise of the noradrenergic and cholinergic systems could be detected in Parkinson’s disease patients with RBD, which was linked to cognitive deficits.^8,9^

Besides deficiencies of neurotransmitters caused by neurodegeneration, differences in the regulation of receptor expression and neurotransmitter metabolism might influence symptom prevalence as proposed for the evolvement of dyskinesias in Parkinson’s disease.^10^ Such receptor-neurotransmitter dysbalance might be particularly relevant for glutamate and its receptors, the brain’s primary excitatory neurotransmitter involved in many physiological processes.^11–14^ Although it has been suggested that glutamate is involved in the pathophysiology of RBD,^15^ its contribution to the clinical phenotype of PD with RBD remains unclear. In addition to ionotropic glutamate receptors (including N-Methyl-d-Aspartate (NMDA), α-Amino-3-hydroxy-5-methyl-4-isoxazolepropionic Acid (AMPA) and kainate receptors), G protein-coupled metabotropic glutamate receptors (mGluR) are paramount for the regulation of glutamate signaling, particularly in the motor circuitry of the basal ganglia.^16^ Specifically, inhibition of mGluR5 can ameliorate motor symptoms and even reduce dopaminergic and noradrenergic degeneration in Parkinson’s disease animal models.^17,18^ Conversely, activation of mGluR5 can lead to amplified neurodegeneration and might reinforce excitotoxicity.^19,20^

Taking together the malignant phenotype of RBD in Parkinson’s disease and the distinctive role of mGluR5 in the basal ganglia motor circuitry and neurodegenerative processes, we aimed to elucidate the relationship between RBD and mGluR5 dysregulation. Using hybrid PET and magnetic resonance (MR) imaging and spectroscopy, we simultaneously examined glutamate metabolite levels and V_T_ of ^11^C-ABP688, a tracer with highly specific binding to the mGluR5, in Parkinson’s disease patients with RBD and compared findings to patients without RBD and healthy control (HC) subjects.

## Materials and methods

### Participants

We recruited 33 patients with Parkinson’s disease, diagnosed according to the current Movement Disorder Society (MDS) clinical diagnostic criteria,^1^ from the tertiary Movement Disorders clinics at the University Hospital Cologne and advertisements in the patients’ magazine of the German Parkinson’s disease association. Twenty-six patients met the criteria for clinically established and seven for probable Parkinson’s disease.^1^ We additionally enrolled 15 HC subjects with no history of movement or psychiatric disorder from newspaper advertisements. Inclusion criteria were age between 51 and 80 years, geriatric depression scale (GDS 15) < 5, and Montreal cognitive assessment (MoCA) score > 22. Exclusion criteria encompassed contraindications for magnetic resonance (MR) imaging or positron emission tomography (PET), known sleep-related breathing disorder, active or former (less than 10 years cessation) smoking as this potentially influences ^11^C-ABP688 binding,^21^ and for Parkinson’s disease patients a disease duration > 15 years. Medication influencing glutamatergic metabolism or signaling (e.g., amantadine and safinamide) had to be stopped five times its individual half-life before MR and PET scanning.

### Clinical assessments

We recorded medical history and current medication in all subjects. Olfactory function was tested with Sniffin’ Sticks, including 12 different odors. Autonomic symptoms were assessed with the scales for outcomes in Parkinson’s disease - autonomic dysfunction (SCOPA-AUT), subjective sleeping quality with the Parkinson’s disease sleep scale (PDSS), and symptoms of RBD with the REM sleep behavior disorder screening questionnaire (RBDSQ). We assessed motor symptom burden using the MDS Unified Parkinson’s Disease Rating Scale III (MDS-UPDRS III) during regular ON state and OFF state 12 hours after discontinuation of dopaminergic medication in patients with Parkinson’s disease. We calculated dopaminergic treatment response as the ratio of the MDS-UPDRS III score during ON state to the score during OFF state. Additionally, we recorded motor symptom duration, Hoehn and Yahr stage, and dopaminergic treatment as levodopa equivalent daily doses (LEDD) according to previously published conversion factors.^22^

### Video-polysomnography

We used a mobile SOMNOscreen™ plus device for overnight video-polysomnography. This device enables 10 EEG recordings (according to the international 10/20 system: F3, F4, C3, C4, O1, O2, A1, A2, Fpz as grounding, and Cz as reference), electrooculography, surface electromyography of the submental muscle and the tibialis anterior muscles, electrocardiography, nasal pressure and thermal flow monitoring, thoracic and abdominal respiratory effort belts, finger pulse oximetry, and synchronized audio-visual recording. Visual PSG scoring was performed by MS, who is a board-certified sleep expert, on 30-second epochs, including total sleep time, sleep efficiency, the absolute amount of stage 1 (N1), stage 2 (N2), slow wave sleep (SWS), and REM sleep, apnea-hypopnea index (AHI, number of apnea plus hypopnea events per hour of sleep), and periodic limb movements in sleep index (PLMSI, number of periodic leg movements per hour of sleep) according to the American Academy of Sleep (AASM) Manual for the Scoring of Sleep and Associated Events Version 2.6.^23^

Diagnosis of RBD was made blinded to clinical and imaging results according to AASM standards by consensus of MS and CD, fellow in sleep medicine for multiple years.^23^ Furthermore, REM sleep without atonia (RSWA) was quantified as the percentage of REM sleep with increased muscle activity of the submental muscle according to the SINBAR criteria using *RBDtector* software (https://github.com/aroethen/RBDtector).^24^ In brief, ‘tonic activity’ was defined as muscle activity persisting > 15s during a 30s REM epoch. ‘Phasic activity’ was scored as brief muscle activity > 0.2s but shorter than 5s, determined considering 3s REM mini epochs. ‘Any activity’ - as the most global measure of RSWA - included ‘tonic’ plus ‘phasic activity’ and muscle activity between 5s - 15s.^24^ All activities are expressed as the percentage of REM sleep affected by RSWA.

### ^11^C-ABP688 PET

Radiosynthesis of ^11^C-ABP688 was performed as previously reported.^25^ All subjects were measured using a Siemens Trio 3T MR scanner with a customized BrainPET insert.^26^ Patients were scanned during a stable ON condition. The average total injected activity per subject was 589.4 ± 25.8 MBq ^11^C-ABP688. Similar to a previous publication,^27^ we applied 50% (range 47.9 - 55.8%) of total activity as a bolus, followed by a 65 min continuous infusion of the remaining 50% (range 44.2 - 52.1%), providing steady-state conditions at 45 - 60 min after bolus injection (Supplementary Figure 1). PET data were acquired in list mode for 65 min and corrected for attenuation using an in-house template,^28^ random and scattered coincidences, decay, and dead time. Image reconstruction was performed using 3D □ OP □ OSEM (2 subsets, 32 iterations), resulting in isotropic voxels of 1.25□mm^3^. We chose a frame scheme with increasing frame duration, including three 5 min frames between 45 - 60 min (Supplementary Figure 1).

We used PMOD 4.0 software and its relevant toolboxes for image analysis. First, dynamic PET images were motion corrected using an averaged image from 5 - 10 min post bolus injection as a template for rigid co-registration. Motion-corrected PET images were rigidly co-aligned to the corresponding anatomical T1-weighted MPRAGE images (TE 2.89 ms, TR 2500 ms, 1 mm^3^ isotropic voxels). Using MR-based segmentation, MR and PET images were normalized to Montreal Neurological Institute (MNI) space to delineate volumes of interest (VOIs) from the built-in Hammers atlas using the PNEURO tool. VOI delineations were manually corrected if necessary, and ^11^C-ABP688 time activity curves from all VOIs were extracted. Bilateral VOIs of the following 13 brain regions were used for further analysis: frontal cortex, temporal cortex, parietal cortex, occipital cortex, insula, anterior cingulate, posterior cingulate, amygdala, hippocampus, caudate, putamen, pallidum, and thalamus. Ascertaining steady-state conditions between 45 - 60 minutes post bolus injection visually (Supplementary Figure 1) and by low coefficients of variance (mean, 1.0 ± 0.6%), we averaged ^11^C-ABP688 concentrations from each VOI during that time.

We also took four plasma samples at 45, 50, 55, and 60 min after bolus injection. In order to obtain metabolite-corrected plasma concentrations, the parent compound was separated from the metabolites in each sample by solid-phase extraction using appropriate cartridges (Waters Sep-Pak® tC18) as previously described.^29^ Again, steady-state conditions of plasma activities during 45 - 60 min post bolus injection were ascertained visually and by a low coefficient of variance (mean, 4.8 ± 2.5%), and we averaged all four metabolite-corrected plasma concentrations. We calculated regional brain V_T_ as follows: The radioligand concentration in the VOI (mean of 45 - 60 min) divided by metabolite-corrected plasma concentration (mean of 45 - 60 min).^30^ Even though total injected activities (*H*(2) = 0.405, *p* = 0.817), plasma concentrations (*F*(2) = 3.193, *p* = 0.050), and parent compounds (*F*(2) = 1.183, *p* = 0.316) did not differ between groups, metabolite-corrected plasma concentrations differed significantly (F(2) = 3.797, *p* = 0.030): HC subjects, 4.1 ± 0.9 kBq/ml, Parkinson’s disease without RBD, 3.5 ± 0.8 kBq/ml, and Parkinson’s disease with RBD, 3.3 ± 1.0 kBq/ml. We, therefore, included metabolite-corrected plasma concentrations as a covariate in our statistical model.

### MR spectroscopy

During PET recording, we simultaneously acquired MR spectroscopy using a single voxel stimulated echo acquisition mode (STEAM) sequence^31^ with the following parameters: echo time (TE) = 6 ms, repetition time (TR) = 4800 ms, mixing time (TM) 47.82 ms, 128 averages, receive-bandwidth = 2000 Hz. Before the acquisition, the radiofrequency power was calibrated for each subject, and B_0_ shim was performed using FASTESTMAP.^32^ The voxel was centered in the left putamen (voxel size: 21 (left-right) × 35 (anterior-posterior) × 21 (rostral-caudal) mm) (Supplementary Figure 2). One extra complete phase cycle was measured without water suppression for eddy-current correction and absolute quantification. All data were preprocessed utilizing the FID-A package in MATLAB 2015a, including removal of motion corrupted scans and phase as well as frequency drift correction.^33^ The metabolite basis set used for quantification in LCModel (6.3-0I) was generated with VeSPA (https://scion.duhs.duke.edu/vespa/)^34^ using previously published chemical shift and J-coupling constants,^35^ including alanine, ascorbate, aspartate, creatine, γ-aminobutyric acid, glucose, glutamine, glutamate, glutathione, glycerophosphorylcholine, myo-inositol, lactate, n-acetylaspartate, n-acetylaspartylglutamate, phosphocreatine, phosphorylcholine, phosphorylethanolamine, scyllo-inositol, and taurine (Supplementary Figure 2). An additional macromolecular spectrum measured using STEAM at 3T obtained from the MM Consensus Data Collection repository (https://mrshub.org/datasets_mm) was also included in the metabolites basis set. For statistical analysis, we only considered the glutamatergic metabolites - glutamate, glutamine, and glutathione - as metabolites of interest in the context of mGluR5 availability.

The anatomical images were segmented using FAST^36^ for cortical grey matter, white matter, and cerebrospinal fluid, and FIRST^37^ for subcortical structures (FMRIB Software Library v6.0.3); the relative amounts within the MR spectroscopy voxel were determined. Metabolite concentrations were corrected for CSF content within the MRS voxel by dividing the concentration value obtained with LCModel by ‘1-CSF fraction’.

Acquisition of MRS failed in three subjects (one subject from each group) due to technical reasons. We additionally excluded three HC subjects, three Parkinson’s disease patients without RBD, and four Parkinson’s disease patients with RBD due to low spectral quality (full-width at half maximum > 0.07 ppm and signal-to-noise ratio < 20).

### Statistical analysis

Data analysis was performed using the Statistical Package for the Social Sciences (SPSS) version 28. Group data are presented as mean ± standard deviation or relative frequencies unless otherwise stated. Normal data distribution was assessed with the Shapiro-Wilk test, Q-Q plots, and box plots. Group comparisons were calculated using Student’s *t*, Mann-Whitney U, and chi-square tests, univariate analyses of variance (ANOVA), analyses of covariance (ANCOVA), and Kruskal-Wallis tests as appropriate. Correlation analyses were calculated using Spearman’s *rho* and Pearson’s *r*, according to the data distribution. VOI-based V_T_ of ^11^C-ABP688 were compared using a repeated measures ANOVA with brain region (n = 13) as within-subject factor and group (n = 3) as between-subject factor. Post-hoc tests were applied to analyze differences in individual brain regions between the three groups. Significance was accepted at *p* < 0.05 uncorrected.

### Standard Protocol Approvals, Registrations, and Patient Consents

The study was approved by the local ethics committee, and written informed consent was obtained from all study participants following the Declaration of Helsinki. Permission to use 11C-ABP688 was obtained from the federal office of radiation safety before the start of the study.

### Data availability

Anonymized data are available upon reasonable request.

## Results

### Clinical assessments

HC subjects (n = 15), patients with Parkinson’s disease without RBD (n = 17), and patients with RBD (n = 16) were comparable in age, sex, cognition, and depressive symptoms (Table 1). Patients with Parkinson’s disease showed reduced olfactory discrimination and reported more autonomic symptoms and a higher burden of sleep disturbances (Table 1). Patients with Parkinson’s disease without RBD and patients with RBD did not differ significantly regarding these non-motor symptoms as well as in clinical metrics of disease burden (e.g., motor symptom duration, motor deficits, and dopaminergic treatment doses, Table 1). Patients with RBD exhibited higher RBDSQ scores than HC subjects and patients without RBD (Table 1).

**Table 1.**
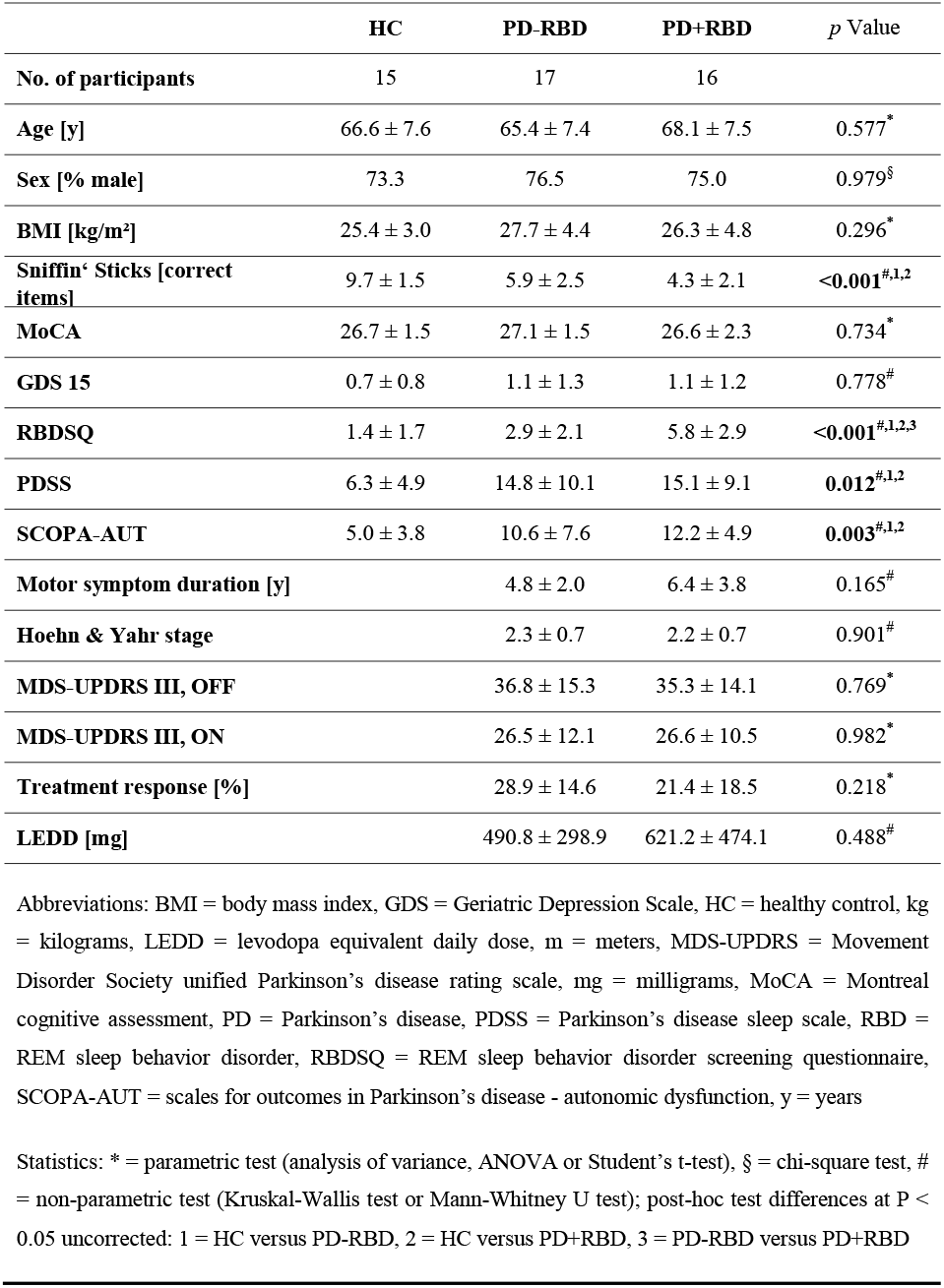
Demographic and clinical characteristics

|  | HC | PD-RBD | PD+RBD | *p* Value |
| --- | --- | --- | --- | --- |
| No. of participants | 15 | 17 | 16 |  |
| Age [y] | 66.6 ± 7.6 | 65.4 ± 7.4 | 68.1 ± 7.5 | 0.577<sup>*</sup> |
| Sex [% male] | 73.3 | 76.5 | 75.0 | 0.979<sup>§</sup> |
| BMI [kg/m<sup>2</sup>] | 25.4 ± 3.0 | 27.7 ± 4.4 | 26.3 ± 4.8 | 0.296<sup>*</sup> |
| Sniffin' Sticks [correct items] | 9.7 ± 1.5 | 5.9 ± 2.5 | 4.3 ± 2.1 | <0.001<sup>#,1,2</sup> |
| MoCA | 26.7 ± 1.5 | 27.1 ± 1.5 | 26.6 ± 2.3 | 0.734<sup>*</sup> |
| GDS 15 | 0.7 ± 0.8 | 1.1 ± 1.3 | 1.1 ± 1.2 | 0.778<sup>#</sup> |
| RBDSQ | 1.4 ± 1.7 | 2.9 ± 2.1 | 5.8 ± 2.9 | <0.001<sup>#,1,2,3</sup> |
| PDSS | 6.3 ± 4.9 | 14.8 ± 10.1 | 15.1 ± 9.1 | 0.012<sup>#,1,2</sup> |
| SCOPA-AUT | 5.0 ± 3.8 | 10.6 ± 7.6 | 12.2 ± 4.9 | 0.003<sup>#,1,2</sup> |
| Motor symptom duration [y] |  | 4.8 ± 2.0 | 6.4 ± 3.8 | 0.165<sup>#</sup> |
| Hoehn & Yahr stage |  | 2.3 ± 0.7 | 2.2 ± 0.7 | 0.901<sup>#</sup> |
| MDS-UPDRS III, OFF |  | 36.8 ± 15.3 | 35.3 ± 14.1 | 0.769<sup>*</sup> |
| MDS-UPDRS III, ON |  | 26.5 ± 12.1 | 26.6 ± 10.5 | 0.982<sup>*</sup> |
| Treatment response [%] |  | 28.9 ± 14.6 | 21.4 ± 18.5 | 0.218<sup>*</sup> |
| LEDD [mg] |  | 490.8 ± 298.9 | 621.2 ± 474.1 | 0.488<sup>#</sup> |

Abbreviations: BMI = body mass index, GDS = Geriatric Depression Scale, HC = healthy control, kg = kilograms, LEDD = levodopa equivalent daily dose, m = meters, MDS-UPDRS = Movement Disorder Society unified Parkinson's disease rating scale, mg = milligrams, MoCA = Montreal cognitive assessment, PD = Parkinson's disease, PDSS = Parkinson's disease sleep scale, RBD = REM sleep behavior disorder, RBDSQ = REM sleep behavior disorder screening questionnaire, SCOPA-AUT = scales for outcomes in Parkinson's disease - autonomic dysfunction, y = years
Statistics: \* = parametric test (analysis of variance, ANOVA or Student's t-test), § = chi-square test, # = non-parametric test (Kruskal-Wallis test or Mann-Whitney U test); post-hoc test differences at $P < 0.05$ uncorrected: 1 = HC versus PD-RBD, 2 = HC versus PD+RBD, 3 = PD-RBD versus PD+RBD

Similarly, polysomnographic metrics of sleep macroarchitecture did not differ between groups, but patients with Parkinson’s disease with RBD exhibited significantly increased tonic, phasic, and any muscle activity during REM sleep compared to patients without RBD and HC subjects (Table 2).

**Table 2.** Polysomnographic metrics

|  | **HC**<br>*n* = 15 | **PD-RBD**<br>*n* = 17 | **PD+RBD**<br>*n* = 16 | *p* Value |
| --- | --- | --- | --- | --- |
| **Sleep period [min]** | 473.1 ± 55.2 | 449.2 ± 74.3 | 455.6 ± 68.4 | 0.609<sup>*</sup> |
| **Sleep efficiency [%]** | 77.4 ± 15.6 | 76.5 ± 21.4 | 84.1 ± 11.4 | 0.428<sup>#</sup> |
| **N1 [min]** | 72.9 ± 47.9 | 62.5 ± 30.3 | 62.1 ± 26.8 | 0.905<sup>#</sup> |
| **N2 [min]** | 173.0 ± 49.9 | 183.5 ± 88.9 | 192.2 ± 51.0 | 0.727<sup>*</sup> |
| **SWS [min]** | 59.0 ± 28.1 | 51.0 ± 34.8 | 53.3 ± 40.5 | 0.807<sup>*</sup> |
| **REM [min]** | 58.1 ± 29.9 | 48.3 ± 32.0 | 73.8 ± 42.0 | 0.122<sup>*</sup> |
| **AHI [/h]** | 12.4 ± 14.1 | 14.5 ± 13.9 | 10.8 ± 11.9 | 0.794<sup>#</sup> |
| **PLMSI [/h]** | 29.0 ± 39.2 | 37.2 ± 47.7 | 26.5 ± 28.2 | 0.817<sup>#</sup> |
| **RSWA, tonic [%]** | 0.5 ± 1.1 | 0.0 ± 0.0 | 11.2 ± 10.5 | <0.001<sup>#,2,3</sup> |
| **RSWA, phasic [%]** | 4.8 ± 3.3 | 4.9 ± 3.0 | 24.6 ± 9.6 | <0.001<sup>#,2,3</sup> |
| **RSWA, any [%]** | 5.2 ± 4.1 | 5.0 ± 3.1 | 35.1 ± 15.5 | <0.001<sup>#,2,3</sup> |

Abbreviations: AHI = apnea-hypopnea index, h = hour, HC = healthy control, min = minutes, N1 = non-rapid eye movement sleep stage 1, N2 = non-rapid eye movement sleep stage 2, PD = Parkinson's disease, PLMSI = periodic leg movements during sleep index, RBD = REM sleep behavior disorder, REM = rapid eye movement sleep stage, RSWA = rapid eye movement sleep without atonia, SWS = slow-wave sleep
Statistics: <sup>\*</sup> = parametric test (analysis of variance, ANOVA), <sup>#</sup> = non-parametric test (Kruskal-Wallis test); post-hoc test differences at *P* < 0.05 uncorrected: <sup>1</sup> = HC versus PD-RBD, <sup>2</sup> = HC versus PD+RBD, <sup>3</sup> = PD-RBD versus PD+RBD

### ^11^C-ABP688 PET and glutamate MR spectroscopy

Repeated measures ANOVA across all three groups revealed significant differences of ^11^C-ABP688 V_T_ (*F*(2,45) = 5.579, *p* = 0.007, Figure 1). Specifically, patients with Parkinson’s disease with RBD showed higher ^11^C-ABP688 V_T_ compared to HC subjects (mean difference 0.534, 95% CI 0.138 – 0.930, *p* = 0.009) and patients without RBD (mean difference 0.580, 95% CI 0.196 – 0.964, *p* = 0.004), whereas the latter two groups did not differ (*p* = 0.812). A significantly higher ^11^C-ABP688 V_T_ in Parkinson’s disease with RBD was observed in all cortical and subcortical brain regions compared to patients without RBD and HC subjects (Figure 2).

**Figure 1:**
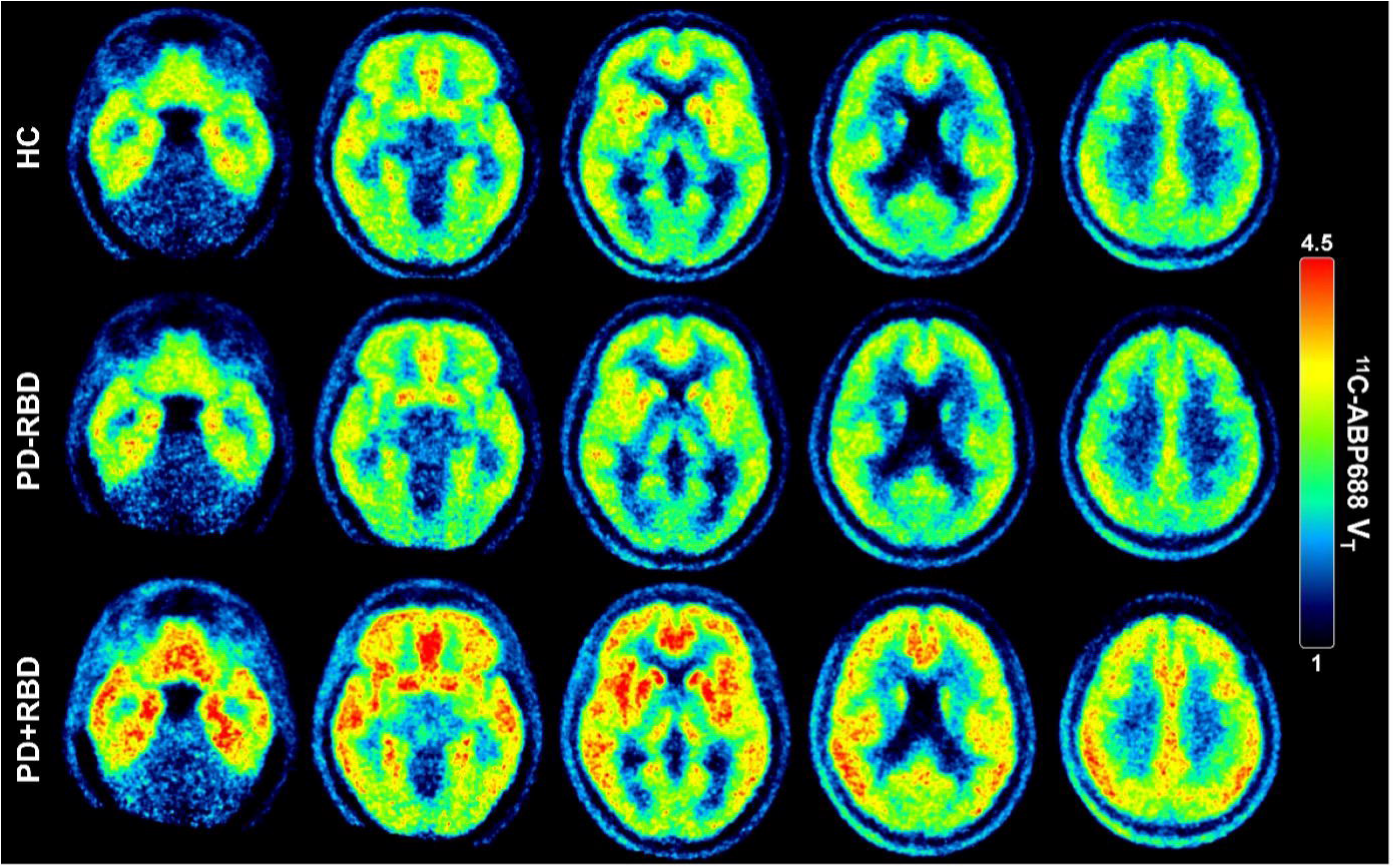
^11^C-ABP688 volumes of distribution (V_T_) images across groups. The top row shows mean V_T_ images sections of healthy control (HC) subjects at different levels, the middle row mean images of patients with Parkinson’s disease without REM sleep behavior disorder (PD-RBD), and the bottom row mean images of patients with Parkinson’s disease with REM sleep behavior disorder (PD+RBD). ^11^C-ABP688 V_T_ images are scaled from 1 to 4.5 ml/cm^3^.

**Figure 2:**
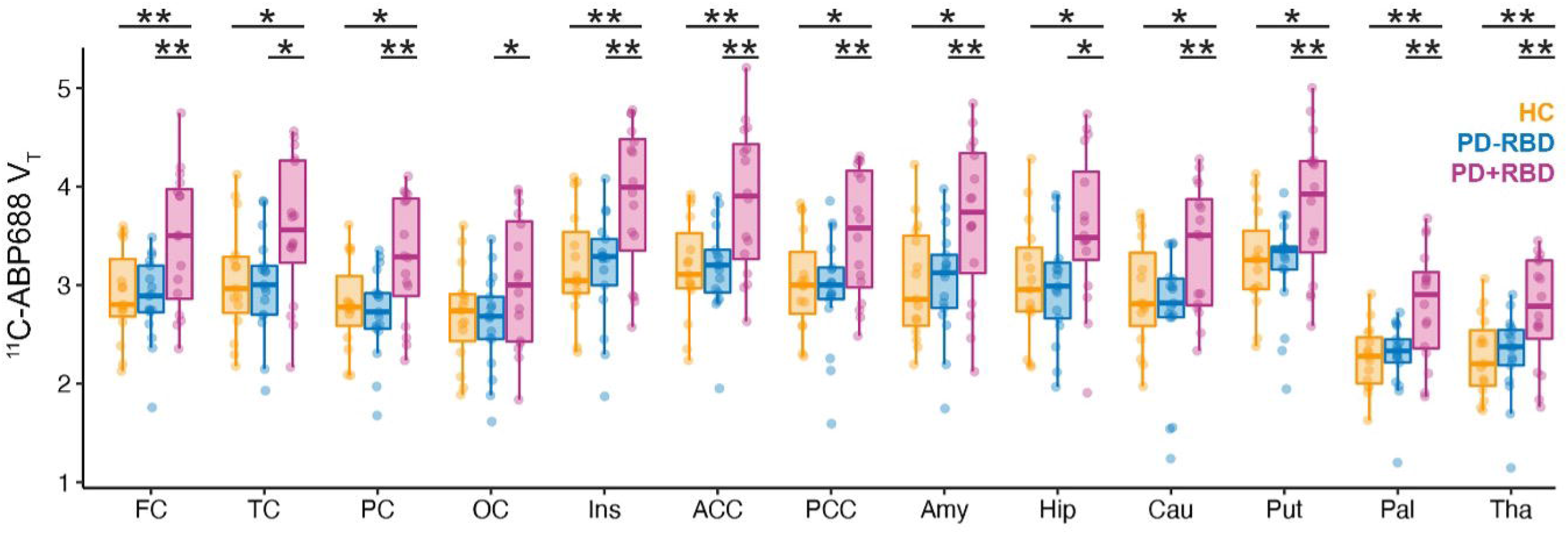
Regional ^11^C-ABP688 volumes of distribution (V_T_) across groups. Boxplots indicate median and upper and lower quartiles; dots represent individual data. Significant differences between groups from *post-hoc* tests of repeated measures ANOVA are indicated by asterisks (*: *p* < 0.05, **: *p* < 0.01, uncorrected). V_T_ is given in ml/cm^3^. Abbreviations: FC = frontal cortex, TC = temporal cortex, PC = parietal cortex, OC = occipital cortex, Ins = insula, ACC = anterior cingulate, PCC = posterior cingulate, Amy = amygdala, Hip = hippocampus, Cau = caudate, Put = putamen, Pal = pallidum, and Tha = thalamus. HC = healthy control, PD-RBD = Parkinson’s disease without RBD, PD+RBD = Parkinson’s disease with RBD.

Group differences remained significant when all demographic and clinical metrics (age, sex, BMI, olfaction, MoCA, GDS 15, RBDSQ, PDSS, and SCOPA-AUT), and metabolite corrected plasma concentrations of ^11^C-ABP688 were considered as covariates (overall effect: *F*(2,35) = 4.787, *p* = 0.015, Parkinson’s disease with RBD versus HC: *p* = 0.047, Parkinson’s disease with RBD versus Parkinson’s disease without RBD; *p* = 0.004).

The comparison of glutamatergic metabolites - glutamate, glutamine, and glutathione - estimated from MR spectroscopy of the left putamen did not reveal significant differences across the three groups (Table 3). The amount of grey and white matter included in the spectroscopy voxel and the spectroscopy quality criteria were comparable across groups (Table 3). None of the estimated glutamate metabolites exhibited a significant covariation with ^11^C-ABP688 V_T_ when evaluated with a repeated measures ANCOVA (all *p* > 0.8, interaction term with brain region: all *p* > 0.2). Additionally, correlation analyses of glutamate metabolites with ^11^C-ABP688 V_T_ of the left putamen did not reveal significant correlations (all |*r*| < 0.100, all *p* > 0.500).

**Table 3.**
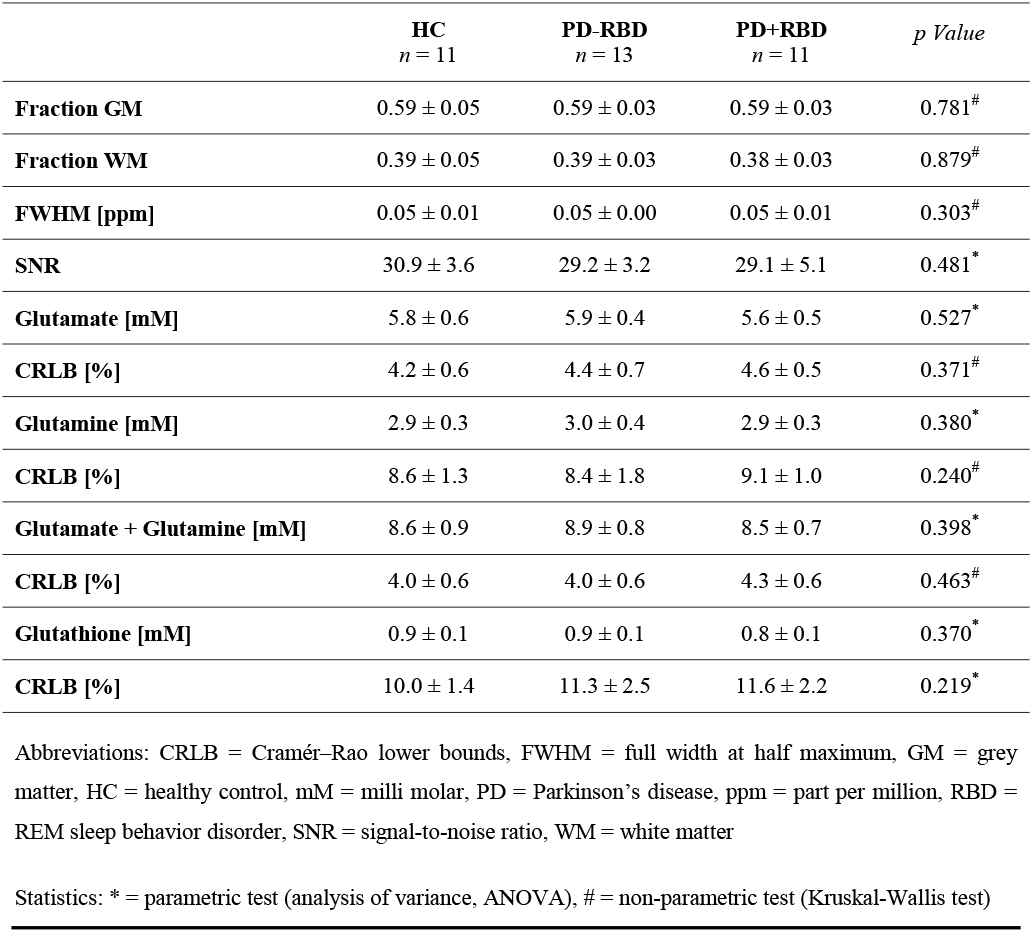
Magnetic resonance spectroscopy estimates of glutamatergic metabolism

### Clinical correlates of ^11^C-ABP688 V_T_

Besides a dichotomized effect of RBD status, we found a linear association between ^11^C-ABP688 V_T_ of all examined brain regions and the amount of RSWA in patients with Parkinson’s disease when including ‘any activity’ in a repeated measures ANCOVA (*F*(1,27) = 4.462, *p* = 0.044, including patients with Parkinson’s disease only and *F*(1,42) = 5.600, *p* = 0.023, including all subjects in analysis).

To interrogate correlates of motor performance in patients with Parkinson’s disease with ^11^C-ABP688 V_T_, we reduced the examined brain regions to critical motor centers (= caudate, putamen, pallidum, and primary motor cortex). Dopaminergic treatment response negatively covaried with ^11^C-ABP688 V_T_ in a repeated measures ANCOVA (*F*(1,30) = 5.823, *p* = 0.022). This was reflected in significant correlations of dopaminergic treatment response with ^11^C-ABP688 V_T_ of the examined motor regions (except caudate) (Figure 3). Motor symptom duration, Hoehn & Yahr stage, and MDS-UPDRS III did not correlate with ^11^C-ABP688 V_T_ of these regions.

**Figure 3:**
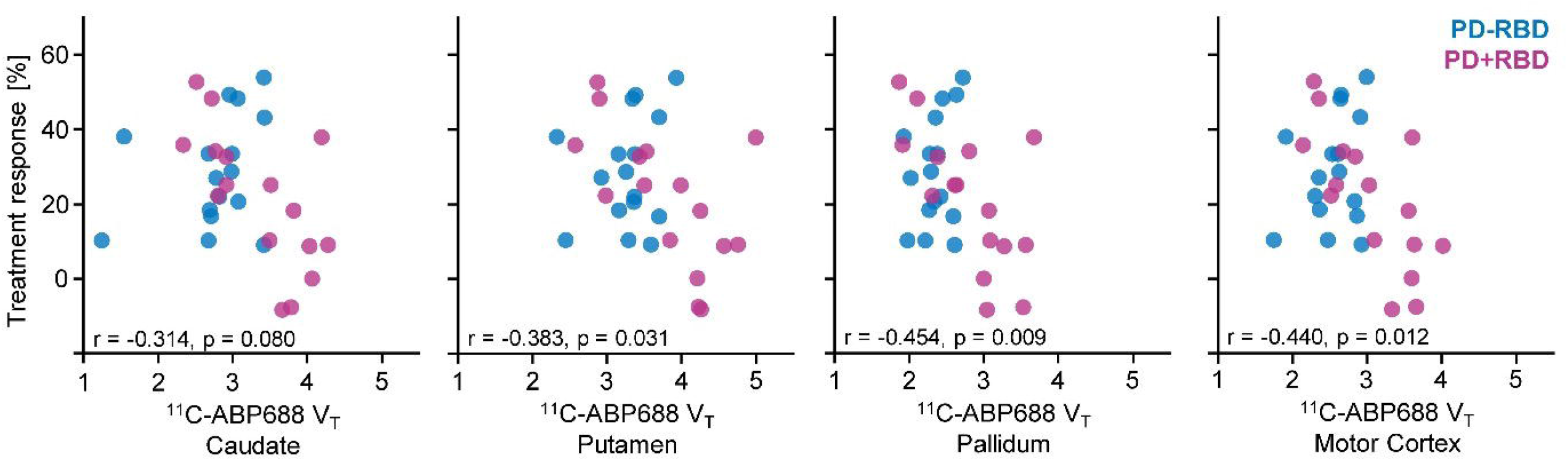
Correlation of ^11^C-ABP688 volumes of distribution (V_T_) with dopaminergic treatment response. Patients with Parkinson’s disease without REM sleep behavior disorder (PD-RBD) are indicated as blue dots and patients with Parkinson’s disease with REM sleep behavior disorder (PD+RBD) in purple. Pearson correlation coefficients and p-values were calculated across all patients. V_T_ is given in ml/cm^3^.

## Discussion

Using simultaneous PET/MR imaging, we discovered higher ^11^C-ABP688 V_T_, but unchanged glutamate, glutamine, and glutathione levels in Parkinson’s disease patients with RBD compared to HC subjects and patients without RBD. We additionally found a linear relationship of increased RSWA as a hallmark of RBD and ^11^C-ABP688 V_T_. Thus, glutamate signaling in Parkinson’s disease patients with RBD seems to be altered at the receptor level despite normal ambient concentrations of glutamate metabolites.

### Glutamatergic neurotransmission in Parkinson’s disease

Most studies assessing glutamatergic neurotransmission in Parkinson’s disease have focused on investigating glutamate, glutamine, or Glx (i.e., the combination of glutamate and glutamine) levels with MR spectroscopy.^38^ In line with our findings, these studies typically did not find significant differences between patients with Parkinson’s disease and healthy controls, even though multiple brain regions, including the substantia nigra,^39^ basal ganglia,^39–43^ thalamus,^44,45^ and cortical regions,^46–49^ were evaluated. As glutamatergic neurotransmission is determined not only by its concentration but equivalently by its receptor availability, our data confirm and extend current knowledge by simultaneously assessing glutamate metabolomics with MR spectroscopy and changes at the receptor level using ^11^C-ABP688 PET, suggesting a dysbalance between these two glutamate signaling factors in Parkinson’s disease patients with RBD.

Interestingly, the few studies that report changes in glutamate levels using MR spectroscopy linked altered glutamate levels with impaired cognition and other neuropsychiatric conditions in Parkinson’s disease.^46,50,51^ These conditions are also associated with RBD in Parkinson’s disease.^52^ As we only included patients without relevant cognitive and psychiatric co-morbidities, changes at the receptor level might be a harbinger of more profound glutamatergic dysregulation in Parkinson’s disease patients with RBD.

### mGluR5, NMDA receptors, and excitotoxicity

mGluR5 (as well as mGluR1) belongs to the Group I of metabotropic glutamate receptors, which are commonly expressed in the perisynaptic zone of the postsynaptic region close to ionotropic glutamate receptors like the NMDA receptor.^53,54^ Upon binding of glutamate, mGluR5 activates Gα_q/11_ proteins, followed by activation of phospholipase C, mobilization of calcium from intracellular storages, and eventually the activation of protein kinase C (PKC).^55^ Additionally, mGluR5 interacts with Homer proteins, connecting them physically to NMDA receptors. mGluR5 contributes to the postsynaptic density, a region in tight apposition to the presynaptic active zone, which is involved in organizing receptors in the synaptic cleft.^56^ Activation of mGluR5 raises the probability of channel opening in NMDA receptors and augments NMDA-mediated currents in neurons.^57,58^ Conversely, activation of NMDA receptors can reverse the desensitization of mGluR5 receptors.^59^ Taken together, these findings indicate that mGluR5 activation augments glutamatergic effects on NMDA receptors, thereby inducing an increased probability of excitotoxicity.^60,61^

In that sense, higher mGluR5 density in Parkinson’s disease patients with RBD could lead to increased activation of NMDA receptors even in the absence of elevated glutamate levels and thereby cause excessive excitatory activity and potentially augment excitotoxicity.^62^ Excitotoxicity is further enhanced by glutamate release from glia cells, which also express mGluR5.^63^ mGluR5 is also reported to be upregulated in reactive astrocytes.^64,65^ However, reduced imidazoline 2 binding sites, a marker of reactive astrocytes, were observed in a recent PET study in mid-stage to advanced patients with Parkinson’s disease compared to HC subjects and early-stage patients.^66^ Hence, as we also examined mid-stage patients, it seems unlikely that the increased ^11^C-ABP688 binding that we observed in Parkinson’s disease patients with RBD is driven by upregulation of the receptor in reactive astrocytes. Instead, we suggest that this change results from dysregulation in neurons.

Notably, activation of mGluR5 can entail a neuroprotective effect in specific cell culture models depending on the stimulation paradigm.^67^ However, the authors conclude that such a switch from facilitation to inhibition of excitotoxicity might more likely occur in the initial phase of an acute event (e.g., as a result of ischemia) but not in chronic neurodegenerative disorders.^67^ Following this notion, excessive glutamatergic action, like the overactivation of NMDA receptors, is more likely to be linked to excitotoxicity, particularly in Parkinson’s disease.^20,68^

Moreover, activation of mGluR5 aggravated 1-methyl-4-phenyl-1,2,3,6-tetrahydropyridine (MPTP)-induced nigro-striatal damage in mice.^19^ Conversely, mGluR5 antagonists showed neuroprotective effects in animal models of Parkinson’s disease, attenuating cell death of dopaminergic neurons of the substantia nigra and reducing microglial activation.^69,70^ Inhibition of mGluR5 in the context of MPTP lesioning also enhanced survival of dopaminergic and noradrenergic neurons in monkeys.^18^

Multiple studies demonstrated that Parkinson’s disease patients with RBD have a faster motor progression and even accelerated dopamine transporter loss and more pronounced brain atrophy compared to patients without RBD, linking RBD in Parkinson’s disease to a diffuse-malignant phenotype of the disease.^6,7,71–73^ Our data indicating a higher mGluR5 density in Parkinson’s disease patients with RBD – potentially leading to amplified excitotoxicity even in the absence of elevated glutamate levels – align with these clinical findings, which suggests a putative pathomechanism. The association between receptor changes and RBD in Parkinson’s disease is strengthened by elevated RSWA, the hallmark of RBD, also being associated with ^11^C-ABP688 binding.

### Association of mGluR5 with dopaminergic treatment

Besides our finding of elevated ^11^C-ABP688 V_T_ in Parkinson’s disease patients with RBD, we could link these changes at the receptor level to reduced dopaminergic treatment effects, which might - in part - provide an additional explanation of a worse motor phenotype in patients with RBD. From animal studies, it is well known that mGluR5 antagonists can mitigate parkinsonian motor symptoms, emphasizing their essential role in the basal ganglia circuitry.^74^ Conversely, mGluR5 activation selectively activated the indirect pathway in a Parkinson’s disease rat model.^75^ More generally, downstream signaling of mGluR5 is supposed to counteract dopaminergic action in the basal ganglia.^16^ Hence, the negative correlation of ^11^C-ABP688 V_T_ and dopaminergic treatment effects we observed in patients with Parkinson’s disease fits well with the animal data.

In the past years, modern mGluR5 antagonists like mavoglurant and dipraglurant were evaluated in phase 2 studies on patients with Parkinson’s disease and revealed good safety and tolerability.^76,77^ However, they failed to show efficacy in reducing levodopa-induced dyskinesia^78,79^ despite substantial pre-clinical data on the beneficial effects of mGluR5 antagonists on levodopa-induced dyskinesias in various animal models of Parkinson’s disease.^80,81^

Our *in-vivo* data hint at a potential role of these drugs as co-medication to facilitate dopaminergic response. Furthermore, their application in Parkinson’s disease patients with RBD might ameliorate the diffuse-malignant phenotype, which warrants further investigation.

### Limitations

We only used a single voxel for MR spectroscopy, centred in the left putamen, to estimate concentrations of glutamate metabolites. We, therefore, cannot exclude concentration differences in other brain regions. Such differences might be detected with more advanced techniques like spectroscopic imaging and chemical exchange saturation transfer (CEST),^82^ which were not used within the constraints of this study. However, we did not observe a significant correlation of the putaminal glutamate metabolites with ^11^C-ABP688 V_T_ in the putamen, and it remains unlikely that global changes of ^11^C-ABP688 V_T_ in Parkinson’s disease with RBD are driven by locally restricted changes in concentrations of glutamate metabolites. Furthermore, MR spectroscopy provides averaged metabolite concentrations of the whole tissue in the voxel but cannot differentiate between the components. We, therefore, analyzed multiple glutamate metabolites including glutamate, glutamine, and glutathione, which are in a tightly interconnected biochemical cycle involving neurons and astroglia, to increase sensitivity to any changes.^83^

We estimated mGluR5 density by ^11^C-ABP688 V_T_, which includes free tracer, specific and non-specific binding of ^11^C-ABP688. We did not calculate more specific non-displaceable binding potentials due to the lack of a suitable reference region for ^11^C-ABP688 devoid of receptors.^84^ However, we thoroughly assessed ^11^C-ABP688 kinetics during the applied bolus-infusion protocol to confirm steady-state conditions for firm quantifications.^30^

We evaluated dopaminergic treatment response in a real-life scenario with quantifying ON motor performance during a regular therapeutic regime and OFF performance after 12 hours of overnight medication withdrawal. Due to ethical considerations, we did not apply sustained withdrawals of medication and standardized doses of levodopa. This might explain why the observed mean treatment response was below 30%, which is demanded by the MDS clinical diagnostic criteria.^1^ However, all patients were additionally interviewed for their subjective treatment response and fulfilled the given diagnostic criteria at the clinically established and probable Parkinson’s disease levels.^1^

## Conclusions

Combining ^11^C-ABP688 PET and MR spectroscopy, we revealed altered glutamate signaling, leading to a dysbalance between changes at the receptor level and glutamate metabolism in Parkinson’s disease patients with RBD. These imaging findings were associated with core clinical features of patients with this malignant phenotype: an increased amount of RSWA and reduced response to dopaminergic treatment. Our data suggest a pathomechanism that might impact the Parkinson’s disease phenotype providing a rationale for further investigations of drugs targeting the mGluR5 in these patients.

## Data Availability

Anonymized data are available upon reasonable request.

## Authors’ contributions

CEJD: Conceptualization, Methodology, Major role in data acquisition, Writing – Review & Editing. AS: Major role in data acquisition, Data processing, Writing – Review & Editing. EF: Methodology, Software, Formal analysis, Writing - Review & Editing. CRB: Methodology, Writing - Review & Editing. LH: Visualization, Writing - Review & Editing. CPF: Formal analysis, Writing - Review & Editing. AG: Methodology, Software, Formal analysis, Writing - Review & Editing. NJS: Resources, Writing - Review & Editing. CWL: Methodology, Software, Resources, Writing - Review & Editing. BN: Resources. KJL: Methodology, Resources, Writing - Review & Editing. GRF: Resources, Writing - Review & Editing, Supervision, Funding acquisition. MS: Conceptualization, Methodology, Formal analysis, Major role in data acquisition, Resources, Data curation, Writing – Original Draft, Visualization, Super vision, Project administration, Funding acquisition.

## Acknowledgments

We thank all study participants and Michael Barbe for his support in patient recruitment. We highly appreciate the technical support of Lutz Tellmann and the professional patient care of Silke Frensch and Suzanne Schaden during PET/MR scanning. We would like to acknowledge E.J. Auerbach and M. Marjanska (Center for Magnetic Resonance Research and Department of Radiology, University of Minnesota, USA) for the development of the STEAM and FASTESTMAP sequences for the Siemens platform, which was provided by the University of Minnesota under a C2P agreement. We also acknowledge D. Deelchand for providing access to the macromolecular spectrum used here.

