## Supplemental Material for "Altered glutamate signaling in Parkinson’s disease patients with REM sleep behavior disorder"

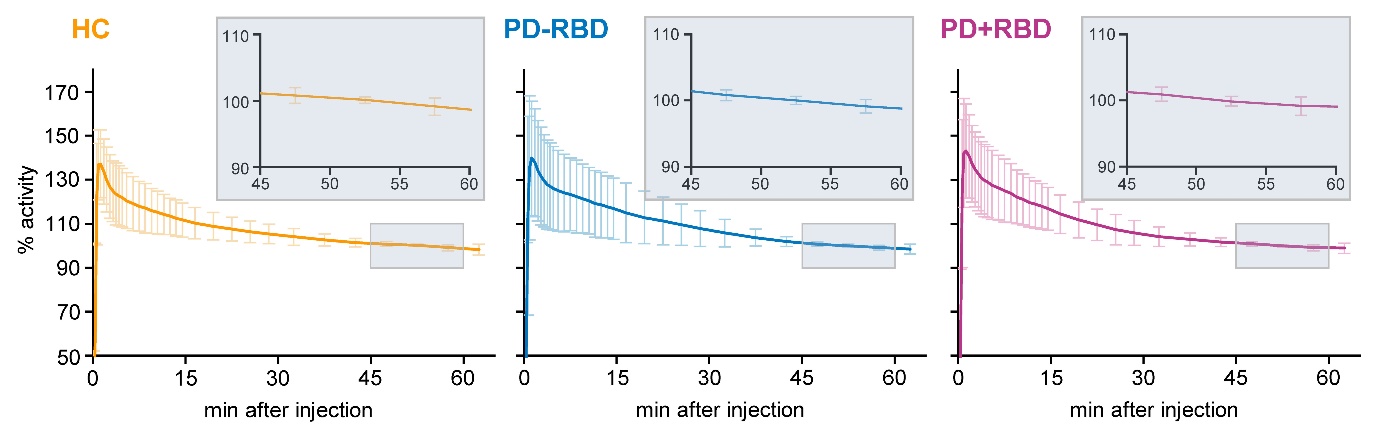

### Additional file 1

**Supplementary Figure 1: ^11^C-ABP688** **time activity curves across groups.** The activity was averaged across all 13 examined brain regions and normalized to the mean activity between 45 - 60 min after bolus injection (set as 100%). Higher activity variability was observed until 30 min after bolus injection as indicated by high standard deviations (vertical bars) during these frames, which was drastically reduced between 45 - 60 min after bolus injection in all groups. Additionally, activity was highly stable, showing less than 2% change between minute 45 and minute 60. Inlets provide magnification of activity during that time.

Abbreviations: HC = healthy controls, PD-RBD = Parkinson’s disease without REM sleep behavior disorder, PD+RBD = Parkinson’s disease with REM sleep behavior disorder

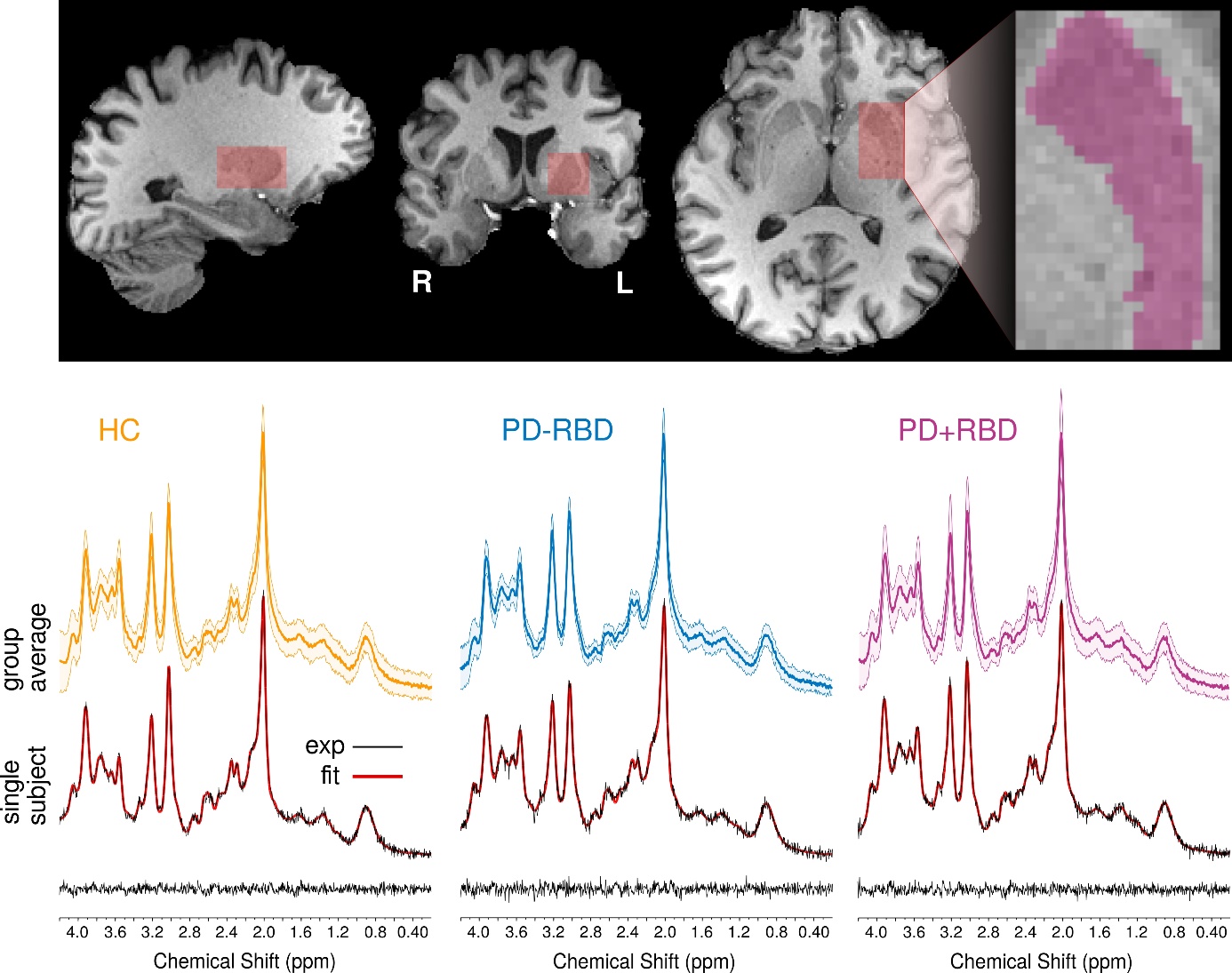

**Supplementary Figure 2: Example of MR spectroscopy placement and resulting spectra.** The upper panel shows a representative example of the voxel placement for MR spectroscopy. The voxel was positioned to cover an applicable extent of the left putamen. The lower panel (top half) provides the group–averaged spectra (light colors indicate standard deviation) for healthy control (HC) subjects (left, yellow), Parkinson’s disease patients without REM sleep behavior disorder (PD-RBD, middle, blue), and Parkinson’s disease patients with REM sleep behavior disorder (PD+RBD, right, purple). Below the group-averaged spectra, representative example spectra for single subjects are given for each group (black lines, experimental data (exp)) together with the LCModel fit (red lines) and the corresponding residuals (bottom).

The metabolite basis set used for quantification in LCModel (6.3-0I) was generated with VeSPA (https://scion.duhs.duke.edu/vespa/) [1] using previously published chemical shift and J-coupling constants, [2] including alanine, ascorbate, aspartate, creatine, γ-aminobutyric acid, glucose, glutamine, glutamate, glutathione, glycerophosphorylcholine, myo-inositol, lactate, n-acetylaspartate, n-acetylaspartylglutamate, phosphocreatine, phosphorylcholine, phosphorylethanolamine, scyllo-inositol, and taurine. An additional macromolecular spectrum measured using STEAM at 3T obtained from the MM Consensus Data Collection repository (https://mrshub.org/datasets_mm) was also included in the metabolites basis set.
